# Prevalence of Dental Caries and Associated Factors, as well as Knowledge, Attitudes, and Practices (KAP) in Children and Adolescents in a Rural Community in Eritrea

**DOI:** 10.1101/2025.03.25.25324603

**Authors:** Meron Tesfay kahsay, Mulue kflom, Ghirmay Ghebrekidan Ghebremeskel, Mengsteab Embaye Gulbet, Huruy Tesfahiwet, Oliver Okoth Achila

**Affiliations:** B. Sc. Clinical laboratory science, Nakfa Hospital, Northern Red Sea branch of the Ministry of Health, Nakfa, Eritrea; Dental technician, Nakfa hospital, Northern red sea branch of the ministry of health, Nakfa, Eritrea; General practitioner, Nakfa Hospital, Northern Red Sea branch of Ministry of the Health, Nakfa, Eritrea; B. Sc. Clinical laboratory science, Eritrean National Health laboratory, Asmara, Eritrea; Unit of Clinical Laboratory Science, Orotta College of Medicine and Health Sciences (OCMHS), Asmara, Eritrea

**Keywords:** Hygiene, Children, Oral health, Prevalence and community health

## Abstract

**Background:** Existing studies in central areas of Eritrea showed high prevalence of dental caries, however, there is gap of knowledge regarding the burden in the rural areas. This study aimed to evaluate the prevalence of dental caries and its associated factors as well as knowledge, attitude and practice of students in Nakfa town, a rural and remote regions of Eritrea.

**Methods:** This study is school based cross sectional study conducted among 401 Junior and high-school students in three schools in Nakfa town. The study used multistage sampling method where finally Students were randomly selected. Self-administered closed end questionnaire with 28 questions were used to collect the data. Dental examination was done by two 5 year experienced and certified dental therapists. All data analysis was done using SPSS version 26.

**Results:** Overall prevalence of dental caries was 65.3% with mean DMFT index and mean significant index (SiC) of 2.03(SD±2.3) and 4.57(SD±1.9), respectively. Mean DMFT index was higher among the older age group (19-23 age group) and those who use traditional stick method of tooth cleaning. Our study also found that higher consumption of porridge was associated with higher prevalence of dental caries as well as higher mean DMFT index. Multivariate analysis showed that age group of 19-23 had 80% higher DMFT index than those 12-14 age category (AIRR= 1.8, 95% CI: 1.1-3). Furthermore, participants who use traditional method of cleaning tooth had 40% higher DMFT than those who use tooth brush with paste (AIRR= 1.40, 95% CI: 1.06-1.8). This study showed taht most of the participants had sufficient knowledge and positive attitude, 61.3% and 73.8% respectively, however, only 8.7% had adequate oral hygiene practice.

**Conclusion:** The prevalence of dental caries was high in Junior and high school students in Nakfa town. High consumption of porridge is associated with high prevalence and high mean DMFT. This study also found that older age group and usage of traditional stick method of teeth cleaning is associated with higher DMFT. The overall level of knowledge and attitude of participants towards oral health was good whereas practice was suboptimal.

## Introduction

Dental caries, also known as tooth decay, is the most common chronic condition to affect children globally. It causes significant negative impacts on the lives of children and young people, including pain, local infection and in some cases may lead to emergency hospitalization due to spread of the infection and systemic illness [1]. In many developing countries, the prevalence of caries has been steadily increasing, largely due to lifestyle changes, the lack of oral health preventive services, and inadequate access to dental care [2]. The growing consumption of sugary food in the developing world, poor tooth brushing habits, poor oral hygiene and low level of awareness about dental caries are some of the factors that increased the levels of Dental Decay [3]. Healthy oral cavity is of great significance for an individual’s overall health and well-being. Further, it enables an individual to masticate, speak and socialize without any active discomfort or embarrassment [4]. The caries process occurs dynamically as bacteria (e.g., Streptococcus mutants) in the mouth utilize free sugars—chiefly sucrose—as an energy substrate. When fermentable carbohydrates are used for energy, pH-lowering acid is produced as a metabolic byproduct. When the critical pH of approximately 5.5 is reached, the tooth enamel and exposed dentin become demineralized. Over time, if this process repeats to the extent and frequency that enamel is not able to sufficiently re-mineralize between acid exposures, caries is formed [5]. Fluoride application can be used to treat early stages of disease; however, many cases require more invasive and expensive restorative treatments (fillings and dental crowns) with the most severe cases requiring extraction. Cases where decay has spread to the pulp of the tooth may also require pulpectomy or root canal therapy [6].

Over the years, there has been reduction in the prevalence of dental caries in the developed nations, probably due to the application of preventive measures [7]. Dental caries is a largely preventable disease, thus there are a range of different programs available to reduce the prevalence in children [8]. One of the barriers to the utilization of preventive dental cares is lack of public awareness and Students are the ideal target group for an early intervention because healthy behaviors and lifestyles developed at younger ages are more sustainable [9,10]. Factors that may be implicated in giving rise to caries in young children have been described in a number of review papers (Federation Dentaire Internationale, 1988; Horowitz, 1998; Moss, 1996; Reisine and Douglass, 1998; Seow, 1998). However, none of these have employed a systematic methodology [11]. Understanding the influences on caries risk knowledge within a country is important for the development of effective and efficient strategies (especially population based prevention strategies) for caries prevention [12]. While consequences for children can be particularly devastating, the progressive nature of dental caries dictates an urgent need for dietary intervention strategies and tools directed at individuals of all ages [13,14].

Nakfa hospital is located in Nakfa district in northern red sea zone and delivers health-care services to residents in the subzone over 50,000. Most of these residents have nomadic way of life in low socioeconomic status. Oral health problem is almost every one’s complaint at least ones in his life, in the region, in which its magnitude and its associated factors is yet to be discovered. Although dental service started in Nakfa hospital in late 2019, it only provides extraction of decayed tooth, which could be prevented by raising the awareness of people towards oral health. Now, our study aimed to assess the prevalence of dental caries and its associated factors. Knowledge, attitude and practices of students towards oral health will also be assessed.

## Methodology

### 3.1 Study design

The study was a questionnaire based cross-sectional survey, conducted amongst 401 junior and secondary school students, in Nakfa town, Northern red sea Eritrea. Multistage random sampling method was used to select the sections for the study. There were 3 junior schools in the town, by cooperating with the ministry Education of the sub zone, two schools (Nakfa Junior school and Smud junior school) were selected randomly. As for the secondary school, the only secondary school (Tsabra junior and secondary school) was selected. After allocation of samples for each school proportional to their population of students, sections were selected randomly from each grade. A simple random method was a choice for selecting students from each section.

### 3.2 Study area

The study was conducted in randomly selected two junior schools and the one secondary school in Nakfa town. As the location of the schools indicate, they can fully represent Nakfa town.

### 3.3 Sample size determination

The sample size determination was based on the level of precision required for the proportion of students who have dental caries, favorable knowledge, attitude and practice towards dental hygiene. There was no documented information on prevalence of caries and favorable KAP towards dental heath in these age category, therefore, we assumed the required precision level to be 50 percent: The sample size is estimated using the Cochran formula.

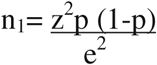

Assumptions for sample size calculation:

p= proportional=0.5 or 50%

z= standard normal distribution (1.96)

e= Margin of error (accepted error) =0.05

n= Targeted sample size from junior school population

The final sample was computed from 1801students, total junior and secondary school students in Nakfa town. Finally, a total of 415 students was targeted for the study considering the discarding rate.

### 3.4 Study population

Students selected by random sampling from each class, with good general health, voluntary to participate and present at the time of data collection was eligible for this study. Additionally, students were told to take consent from their family before one day of data collection. Those students with severe general health problems that affect oral health, with dental emergencies and not voluntary were excluded.

### 3.5 Questionnaire preparation

The format of questionnaire was adapted from questionnaire used in the study by Anshu.B et.al [4] with some modifications. The pretested self-administered questionnaire containing 28 questions was translated in to local language Tigre by collaborating with experts in both English and Tigre languages. Questions was divided into five domains addressing demographics, knowledge, attitudes, practice and dietary history. The questionnaire comprises of 11 questions to access to their knowledge, 7 attitude related questions, 11 practice related questions and 4 dietary related questions. Pre-testing of the questionnaire was carried out by initially administering the questionnaire to 10 students. The questionnaire was then modified accordingly and administered to all participants.

The first section questions were information regarding the socio-demographic background of the participants. In the next section participant’s knowledge was gathered about oral hygiene with total of 10 questions. For assessing knowledge, correct answers were given a score of one whereas incorrect answers and ‘I don’t know’ answers were given a score of zero. Participants who scored 6 or more (>60%) were categorized as belonging to high knowledge group and those with a score of five and below were considered to be in low knowledge group.

Section three consisted of seven questions exploring the attitude and perception of the participants regarding oral health. Positive attitude responses were given a score of 1 and negative responses were given a score of negative one (-1) and ‘I don’t know response’ were given zero score. Those who scored four or more were considered to be having a positive attitude and those scoring less than four were considered having a negative attitude towards oral health. The fourth section assessed the participant’s oral health practices, with total of 11 questions. Correct answers were given a score of one whereas incorrect answer and ‘I don’t know’ answer were given a score of zero. Participants who answered seven and more questions were considered as having adequate practice, whereas, with less than seven correct answers were not adequate practice. The dietary questions were comprised of 4 questions.

### 3.8 Survey methodology

Informed consent was obtained from each participant before the administration of the questionnaire, explaining the need and the purpose of the study. The response format included multiple choice questions in which the students were instructed to choose one or more from the provided list of options. The students were then given instructions regarding filling the questionnaire. Furthermore, the investigator was present while the questionnaire was being filled and all queries of participants was addressed by the investigators. The students were informed to fill in the questionnaire without discussion with each other within half an hour time.

Dental caries experience among the participants was assessed using the Decayed, Missing and Filled Teeth (DMFT) index, which is a measure of life time dental caries experience in permanent dentition. Clinical oral examination of the students was performed by qualified dental therapists one from Nakfa hospital and one from Afeabet hospital following World Health Organization (WHO) diagnostic criteria. All examinations were done in the classrooms with the students seated in an upright chair.

### 3.9 Statistical analysis

The collected questionnaire was checked for its’ completeness and consistency first, it was entered in to CS pro version 7.4 software and it was organized, robustly cleaned and analyzed using statistical package for social science(SPSS) version 26. Comparison of numeric variables was done by Mann-Whitney u Test and Kruskal Wallis test. Comparison of nominal variables was done by Pearson chi square test. Correlation of two numeric variables was analyzed using spearman’s correlation coefficient.The negative binomial regression model was used to examine the factors associated with DMFT, adjusting for other independent variables. An incidence rate ratio (IRR) with a 95% confdence interval (CI) was utilized to express the magnitude of the association between the independent and dependent variables. P values less than 0.05 was considered signifcant.

### 3.10 Operational definition

**Dental caries.** The presence of tooth decay, missing and filled teeth at the time of oral examination.

**Decayed tooth (D).** Includes carious teeth, filled teeth with recurrent decay, a tooth with the only root left, temporarily filled teeth surface with other surfaces (parts) decayed.

**Missed tooth (M).** Includes tooth that is missed due to caries but it doesn’t include teeth missing for reasons other than caries.

**Filled teeth (F).** Includes teeth that have one or more permanent restoration with no secondary (recurrent) caries.

**First quadrant tooth:** Upper right side from midline

**Second quadrant tooth:** Upper left side from midline

**Third quadrant teeth:** Lower left side from midline

**Fourth quadrant:** Lower right side from midline

**The DMFT index:** is the average number of teeth per person that are decayed (D), missed because of caries (M), or filled (F). It is a quantitative expression of the lifetime caries experience of the permanent teeth. In the calculation of the DMFT index, the numerator is the sum total of DMF teeth and the denominator is the total number of persons examined [15].

**Mean Significant Index(SiC):** The SiC index is the mean DMFT of the top third of the study group with the highest caries score [21].

#### Patient and public involvement

The patients and public were not involved in any way in the design, or conduct, or reporting, or dissemination plans of the research.

## Result

### Sociodemographic characteristics of the participants and association of oral hygiene practice of participants with gender

Total of 415 data were collected and 14 questionnaires were discarded due to incomplete data. Finally, data of 401 participants were subjected to data analysis. The average age of participants was 16, with minimum and maximum ages 12 and 23 respectively. Males hold 53.6% of the study participants and females were 46.4%. Most of the participants were in the age range of 15-18(65.6%) followed by 12-14 (27.7%) and 19-23(6.7%) age category, consecutively. Junior students were 44.6% and high-school students were 55.4%. Toothbrush were mostly used by females than males, 51.1% and 18.6% respectively. Even though, males use dominantly traditional stick to clean their teeth they do it for longer time duration than females,72.8% and 60.8% respectively. Majority of the participants (79.3%) rinse their teeth after ever meal with no significant difference seen in males and females. Higher proportion of females use dentist directed teeth cleaning method than males. There was no single participant who attended regular dental checkup (every 6 months) in this study. As the data of participants who use toothbrush alone analyzed, higher males proportion used pea size toothpaste than females, which is the recommended one (table-1).

**Table-1.** Overview of participants sociodemographic and Oral hygiene habits of participants across gender.

| Variables | category | sex |  | p-value | Total count(%) |
| --- | --- | --- | --- | --- | --- |
|  |  | Male count(%) | Female count(%) |  |  |
| Age category | 12-14 | 47(21.9) | 64(34.4) | **0.001** | 111(27.7) |
|  | 15-18 | 146(67.9) | 117(62.9) |  | 263(65.6) |
|  | 19-23 | 22(10.2) | 5(2.7) |  | 27(6.7) |
| Grade category | Junior | 75(35) | 104(55.9) | **<0.01** | 179(44.8) |
|  | High-school | 139(65) | 82(44.1) |  | 221(55.2) |
| Oral hygiene method used | Toothbrush and paste | 40(18.6) | 95(51.1) | **<0.01** | 135(33.7) |
|  | Other than toothbrush and paste | 171(79.5) | 90(48.4) |  | 261(65.1) |
| Frequency of tooth cleaning | Twice and more a day | 84(39.6) | 74(40) | 0.939 | 158(39.4) |
|  | Less than twice a day | 128(60.4) | 111(60) |  | 243(60.5) |
| Mouth cleaning interval | morning before breakfast and night before going to bed at least | 56(26.3) | 45(24.2) | 0.63 | 101(25.1) |
|  | Other time | 157(73.7) | 141(75.8) |  | 298(74.3) |
| Duration of brushing | Two and more minutes | 155(72.8) | 113(60.8) | **0.001** | 268(67.2) |
|  | Less than two minutes | 13(6.1) | 33(17.7) |  | 46(11.5) |
|  | Do not know | 45(21.1) | 40(21.5) |  | 85(21.3) |
| Mouth rinsing after every meal | Yes | 169(79.3) | 149(80.1) | 0.73 | 318(79.7) |
|  | No | 20(9.4) | 20(10.8) |  | 40(10) |
|  | Do not know | 24(11.3) | 17(9.1) |  | 41(10.3) |
| Brushing technique used ? | As directed by dentist | 17(7.9) | 32(17.2) | **0.005** | 49(12.2) |
|  | Not directed by the dentist | 198(92.1) | 154(82.8) |  | 352(87.8) |
| Toothpaste used for those who use toothbrush? | Fluoridated | 22(55) | 62(65.3) | 0.52 | 84(62.2) |
|  | Herbal | 4(10) | 8(8.4) |  | 12(8.9) |
|  | Do not know | 14(35) | 25(26.3) |  | 39(28.9) |
| Amount of toothpaste used for toothbrush users? | Pea size | 12(30) | 9(9.5) | **0.003** | 21(15.6) |
|  | Other than pea size | 28(70) | 86(90.5) |  | 114(84.4) |
| Consumption of biscuits and creams | At least once a day | 90(41.9) | 90(48.4) | 0.3 | 180(44.9) |
|  | At least once a week | 99(46) | 74(39.8) |  | 173(43.1) |
|  | Never | 26(12.1) | 22(11.8) |  | 48(12) |
| Consumption of tea or coffee with sugar | At least once a day | 165(76.7) | 147(79) | 0.19 | 312(77.8) |
|  | At least once a week | 37(17.2) | 22(11.8) |  | 59(14.7) |
|  | Never | 13(6) | 17(9.1) |  | 30(7.5) |
| Consumption of porridge | At least once a day | 71(33) | 60(32.3) | 0.23 | 131(32.7) |
|  | At least once a week | 116(54) | 111(59.7) |  | 227(56.6) |
|  | Never | 28(13) | 15(8.1) |  | 43(10.7) |
| Consumption of candy | At least once a day | 108(50.2) | 108(58.1) | **0.017** | 216(53.9) |
|  | At least once a week | 100(46.5) | 64(34.4) |  | 164(40.9) |
|  | Never | 7(3.3) | 14(7.5) |  | 21(5.2) |

### Prevalence of dental caries in Nakfa town and association of dental caries with practice habits of participants towards oral health,2022

Overall prevalence of dental caries among the participants was 65.3% with mean DMFT index and mean Significant index (SiC) was 2.03±2.3 and 4.66±2.2 respectively. From this, decayed accounts for 96.5% of the total DMFT index with mean decayed teeth of 1.96±2.3 and missed teeth accounts for the rest with mean missed teeth of 0.07±0.28, there was no participant with filled teeth. Third and fourth quadrants were the most affected gum regions, with mean DMFT of 0.7±0.8 and 0.67±0.8 respectively, while first and second quadrants had mean DMFT of 0.34±0.65 and 0.67. Mean DMFT index was higher among the older age group and those who use traditional stick method of tooth cleaning. Prevalence of dental caries was also higher among the older age group with close to significance, however, method of teeth cleaning had no effect in prevalence of dental caries. The participants with higher consumption of porridge had higher prevalence of dental caries and higher mean DMFT (table-2).

**Table-2.**
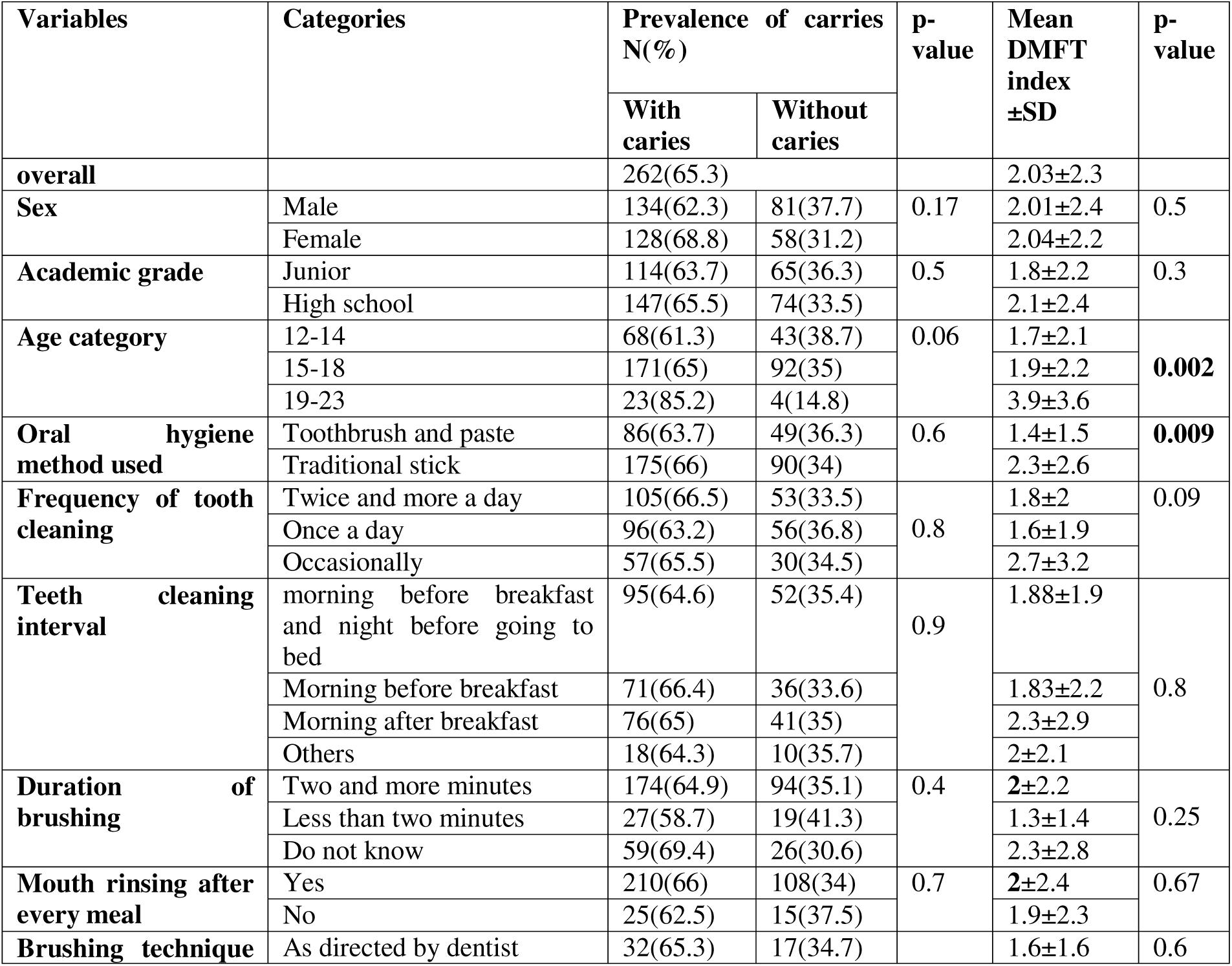
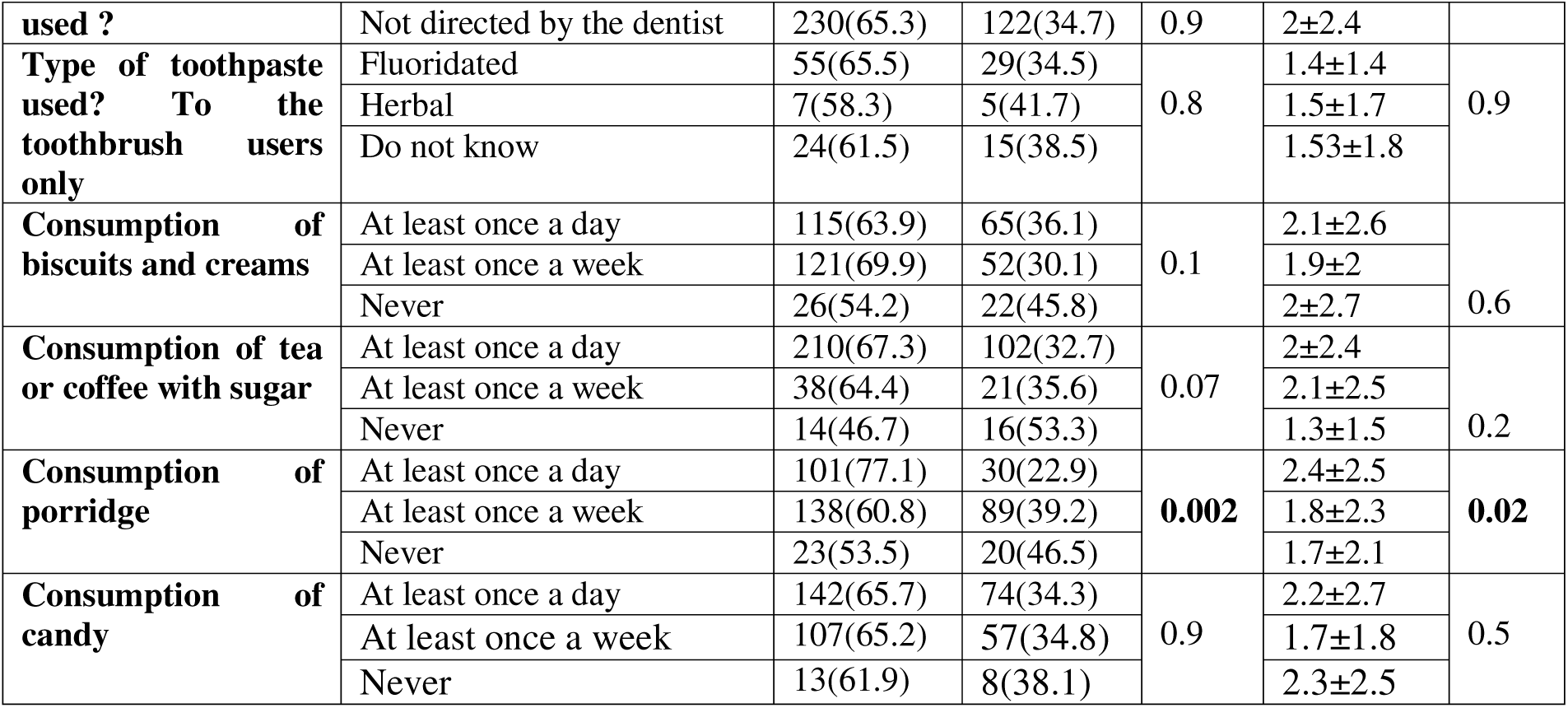
prevalence of dental caries and its associated factors among junior and high-school students of Nakfa town,2022.

Multivariate analysis showed that age group of 19 up to 23 had 80% higher number of decayed and missed teeth than those 12 up to 14 age category. And those who use traditional method of cleaning tooth had 40% higher number of dmft than those who use toothpaste and tooth brush (table-3).

**Table-3.**
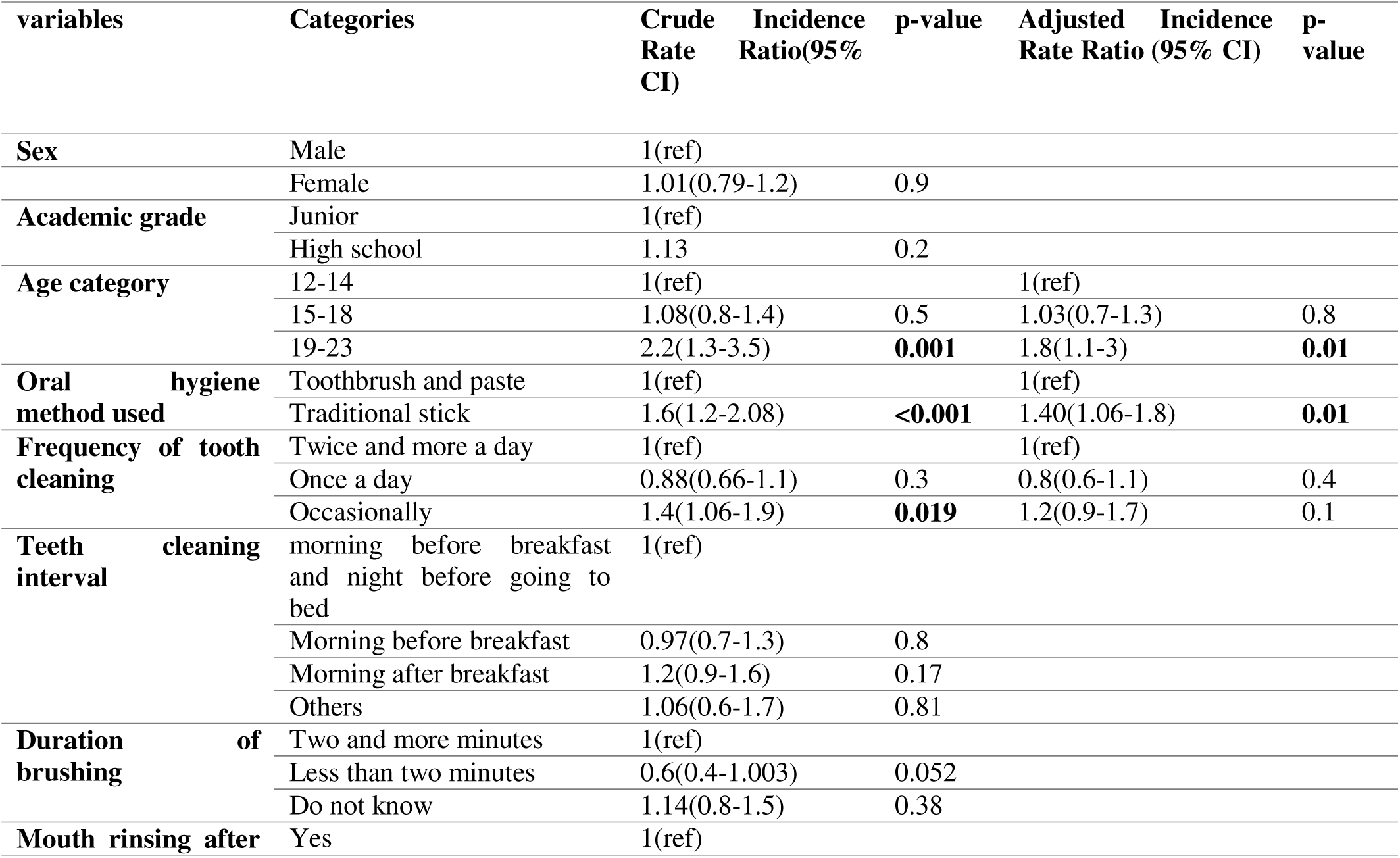
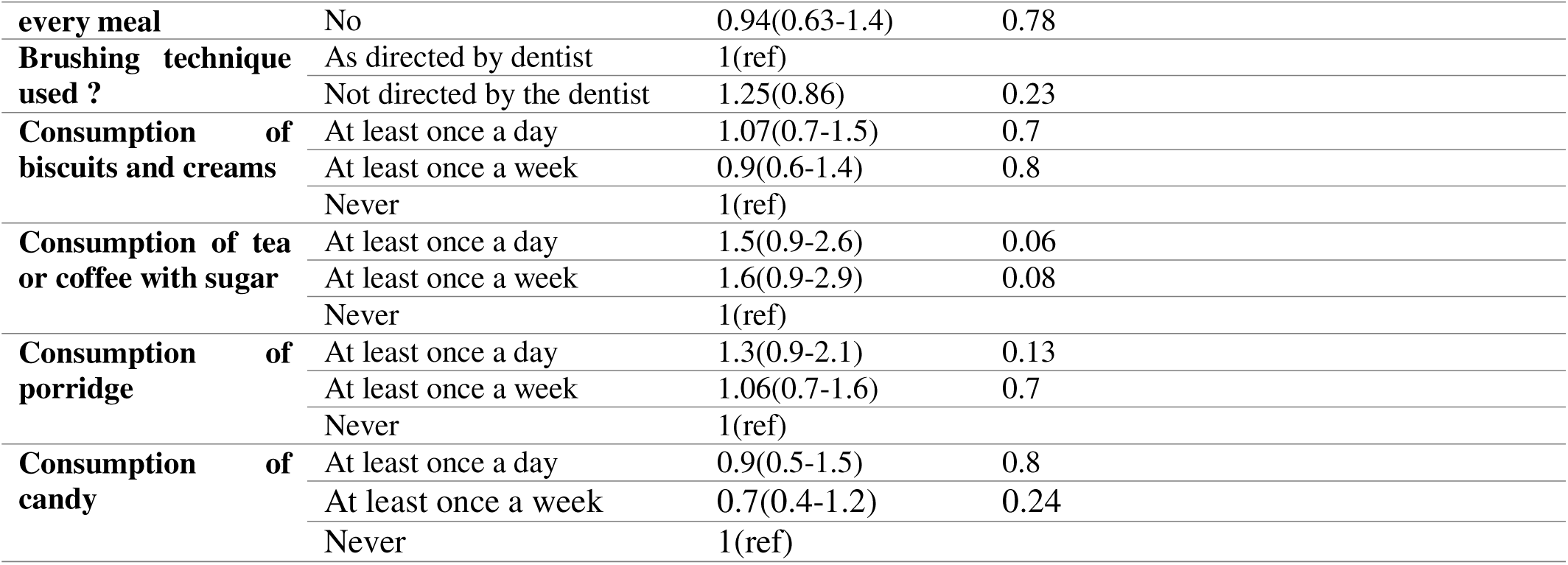
Negative binomial regression: Factors associated with number of dmft.

## Respondents knowledge on oral health

In regard the knowledge of the participants, interestingly minor students believe any food can cause dental caries. However, most of the participants believe that fluoride in tooth paste makes teeth resistant to carries and also reported that sweet, smoking, consumption of soft drinks, alcohol consumption and paan/tobacco chewing affects’ dental health negatively. Apart from this, only about quarter of them know that gum bleeding and tenderness is sign of gum inflammation and few know that dental plaque means soft deposit in teeth. The consequence of infrequent brushing was answered correctly by majority i.e. staining of teeth, dental plaque, bleeding from gum and dental caries. Regarding the overall knowledge of the participants, 61 % had sufficient knowledge (Table-4).

**Table-4.**
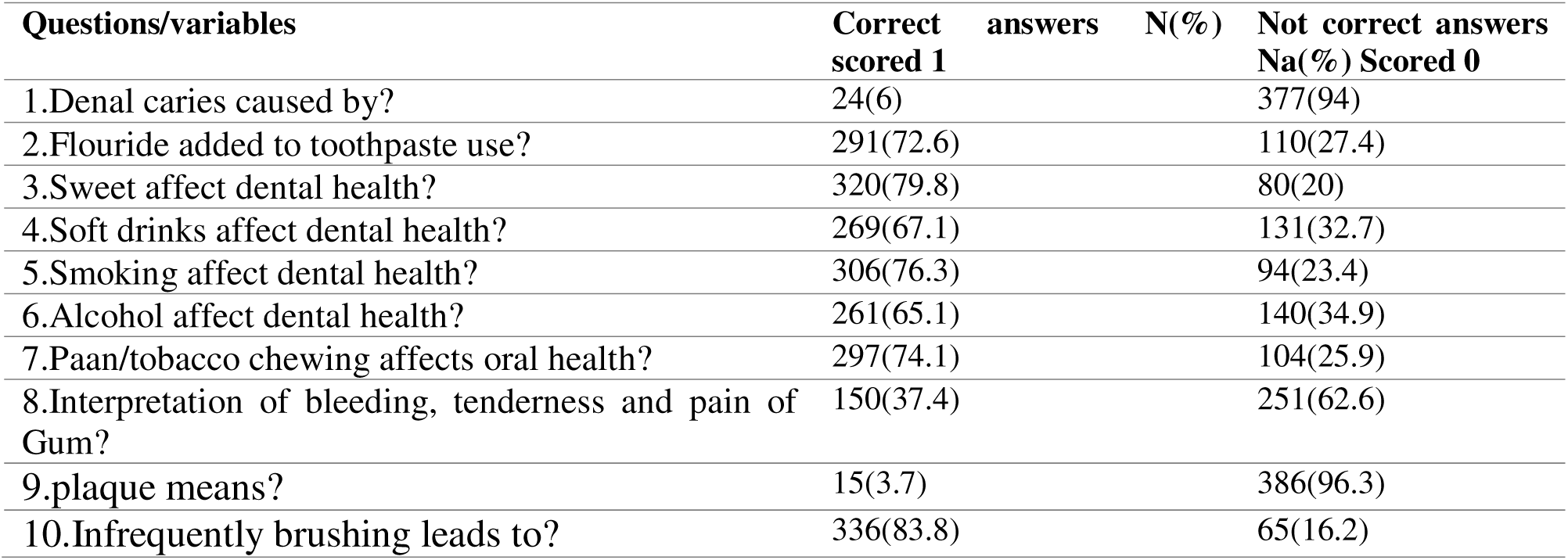
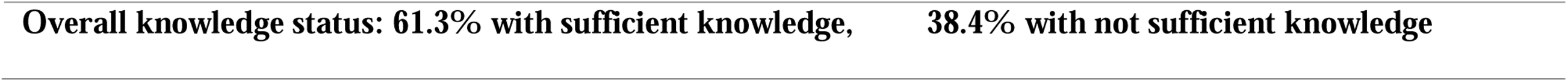
Response of the participants to knowledge and awareness questions.

## Respondents Attitude towards oral health

The response of participants towards attitude questions was found that: majority’s perception was dental caries affects’ aesthetics and general body health and also believe tooth decay is avoidable. Moreover, great portion of the subjects said that care of teeth and regular dental checkup is necessary. 60% of the participants had attitude that maligned teeth affect dental health and also most of them believe choosing the right tooth brush is important. Overall, 73% of the participants had positive attitude towards oral health (table-5).

**Table-5.** Response of the participants to Attitude questions.

| Questions/variables | Positive attitude<br>N(%) scored 1 | Neutral attitude<br>N(%) scored 0 | Negative attitude<br>N(%) scored -1 |
| --- | --- | --- | --- |
| 1. Do you think caries affect aesthetics? | 224(55.9) | 85(21.2) | 91(22.7) |
| 2.Do you think cries affect general health? | 393(98) | 4(1) | 3(0.7) |
| 3.Do you think tooth decay is avoidable ? | 288(71.8) | 56(14) | 57(14.2) |
| 4.Do you think care of teeth necessary? | 330(82.3) | 51(12.7) | 20(5) |
| 5.Do you think maligned teeth affect oral health? | 242(60.3) | 92(22.9) | 67(16.7) |
| 6.Do you think regular dental checkup is necessary? | 330(82.3) | 53(13.2) | 18(4.5) |
| 7.Do you think it is important to choose right tooth brush? | 334(83.3) | 43(10.7) | 24(6) |
| Overall attitude status:73.8% with positive attitude , 25.9% with negative attitude |  |  |  |

Our finding reveals, most of the participants did not used tooth brush with paste, they did not clean their teeth twice and more daily and brushing interval they accustomed to use was other than morning before breakfast and night before going bed. Moreover, majority of students either did not use tooth paste or they used toothpaste without fluoride and also they did not used pea size tooth paste. Regarding bristle type, most of them used other than soft bristle and only third of the participants replaced their toothbrush at least every three months. Minority used brushing technique requested by dentist and none of the participants attended regular dental checkup i.e. every 6-12 month. Although, majority used traditional way of cleaning teeth, they clean their teeth for two or more minutes and majority they rinse their teeth after every meal. Regarding overall practice status of participants, only 8.7% of the participants scored good practice behavior (Table-6).

**Table-6.** Response of the participants to the practice questions.

| Practice questions | Correct answers<br>N(%) scored 1 | Not correct answers<br>N(%)scored 0 |
| --- | --- | --- |
| 1.Oral hygiene method used? | 135(33.7) | 266(66.3) |
| 2.Frequency of tooth brushing? | 158(39.4) | 239(59.6) |
| 3.Brushing intervals? | 101(25.2) | 298(74.3) |
| 4.Duration of brushing? | 268(66.8) | 131(32.7) |
| 5.Toothpaste used? | 84(20.9) | 317(79.1) |
| 6.Amount of tooth paste used? | 21(5.2) | 380(94.8) |
| 7.Type of tooth brush used? | 41(10.2) | 360(89.8) |
| 8.Tooth brush replace after? | 129(32.2) | 272(67.8) |
| 9. Mouth rinsed after every meal? | 318(79.3) | 81(20.2) |
| 10.Brushing technique used? | 49(12.2) | 352(87.8) |
| 11.Frequency of dental visit? | 0(0) | 401(100) |
| Overall practice status: 8.7% with adequate practice , 90.5% with not adequate practice |  |  |

There was no significant difference seen between male and female in regard of their overall knowledge, attitude or Practice in our study. Surprisingly, students with adequate practice were higher in junior school than high-school (p-value, 0.01), but there was no significant difference seen between them in respect of attitude and knowledge (table-7).

**Table-7.** knowledge, Attitude and practice association with variables.

| Variables |  | Sex |  |  | Academic grade |  |  |
| --- | --- | --- | --- | --- | --- | --- | --- |
|  |  | Male<br>count(%) | Female<br>count(%) | p-value | Junior<br>Count(%) | High-school<br>count(%) | p-value |
| **Knowledge** | High | 127(59.3) | 119(64) | 0.34 | 107(59.8) | 139(63.2) | 0.48<sup>p</sup> |
|  | Poor | 87(40.7) | 67(36) |  | 72(40.2) | 81(36.8) |  |
| **Attitude** | Positive | 154(72) | 142(76.3) | 0.31 | 132(73.7) | 164(74.5) | 0.85<sup>p</sup> |
|  | Negative | 60(28) | 44(23.7) |  | 47(26.3) | 56(25.5) |  |
| **Practice** | Adequate | 14(6.6) | 21(11.4) | 0.093 | 23(12.8) | 12(5.5) | **0.01<sup>p</sup>** |
|  | Inadequate | 199(93.4) | 164(88.6) |  | 156(87.2) | 207(94.5) |  |

p: pearson chi square test

Interpretation of correlation was adopted from pragya.p.w et.al [16]. R value (**r**) of 0-0.25= weak correlation, between 0.25 and 0.5=fair correlation, 0.5-0.75= good correlation and >0.75=excellent correlation. The correlation of knowledge with attitude was fair, knowledge with practice had also fair correlation and attitude with practice showed weak correlation (table-8).

**Table-8.** correlation of knowledge, Attitude and practice.

| Variable | Correlation coefficient | p-value |
| --- | --- | --- |
| **Knowledge-attitude** | 0.414<sup>s</sup> | <0.01 |
| **Knowledge-Practice** | 0.279<sup>s</sup> | <0.01 |
| **Attitude-Practice** | 0.146<sup>s</sup> | <0.01 |

S: spearman's correlation

## Discussion

Regarding the use of oral hygiene practices across gender, this study reveals toothbrush was mostly used by females than males, besides, higher proportion of females use dentist directed teeth cleaning method than males. This can be attributed to the fact that females care more about their aesthetics than males, this is consistent with other studies with similar findings [30,31]. Our finding also shows males clean their teeth for longer time duration than females, this is plausible when we consider higher proportion of males use traditional stick method for cleaning their teeth, in which people use it for longer period traditionally. Majority of the participants rinse their teeth after ever meal, which is a good culture among the population that enforces every person to respect his/her hygiene before every prayer locally known as “Selat”. Regular dental checkup was attended by none of participants in the study due to probably lack of awareness of population towards dental clinic due to short duration since establishment of dental clinic in 2019.

The overall prevalence of caries among study participants was 65.3%. This was slightly lower than the previously done studies in Asmara; Nada.A et.al (67.9%) and Amanuel.k et.al (78%) [20,21], but the trend showed that prevalence is increasing as compared to two decades ago i.e 50% [22]. As we compare this with results from sub-Saharan countries, Kenya (43.3%) [23], Ethiopia (48.5%) [29] and Nigeria (55%) [24], this result is higher. The overall mean DMFT index of the study participants was 2.03(SD 2.3), this is included in the low prevalence group in the WHO classification [25]. This was also lower than other studies done in Asmara [20,21]. In the urban areas of Eritrea people do not use commonly traditional stick for teeth cleaning due to shortage, however, in rural areas like Nakfa region it is common, this could be the reason for lower DMFT in Nakfa town. The rare usage of traditional way(stick) of cleaning teeth in the urban areas of Eritrea could make this difference visible. However, this was still higher than studies done in Kenya (0.42) [26], Ethiopia (1.3) [20] and Sudan (0.42) [27]. The third and fourth quadrants were the most affected teeth parts, this can be explained by the higher effect of gravity on them. The overall mean significant index(SiC) was 4.57±1.9, this was higher than the WHO suggestion that every nation’s SiC value to be below 3 by 2015 [28], but it is comparable with study done in Asmara [21]. Our study founds Mean DMFT index was higher among the older age group (19-23 age group), this was consistent with study done by Nada A. et.al in which there was a very strong correlation between age and mean DMFT (r = 0.91). This study reveals method of teeth cleaning had no effect in prevalence of dental caries, however, those who use traditional stick method was associated with higher mean DMFT index than those who use tooth brush. The participants with higher consumption of porridge had higher prevalence of dental caries and higher mean DMFT. The high carbohydrate content of porridge and its sticky nature could be the main reason for this.

Regarding the multivariate analysis; only higher age group and use of traditional stick for cleaning teeth was associated with higher number of dmft. In our study generally consumption of sweet foods or drinks did not associate with higher number of dmft, this was in discordant with other studies in which intake of dietary sugars is the most important risk factor for the development of caries [32,34].

This study revealed that majority of the students missed that any food can cause dental caries. Majority (65.3%) believes only sweet causes dental caries implying that they could be careless about cleaning their teeth after eating any food, this can support the high prevalence of dental caries. This study also showed most of the participants know the use of fluoride in toothpaste, this was coherent with study done in north eastern India which also revealed more than half of the participants were aware about the role of fluoride in preventing dental decay [16]. Contrary to this, other studies in Saudi and Nepal students [17,18], showed fewer students were with similar awareness. Our finding also shows that most of the participants believe; sweet, smoking, consumption of soft drinks, alcohol intake and paan/tobacco chewing affects’ dental health negatively. Study done by Nshu B. et.al also reported similar finding [4]. Pragya.p et.al founds that 91.7% knew the ill effects of tobacco; however, 34.4% still indulged in paan chewing which is noteworthy [16]. Apart from this, only about quarter of them know that gum bleeding with tenderness is sign of inflammation and few know that dental plaque means soft deposit in teeth. Regarding the overall knowledge of the participants, 61 % had sufficient knowledge.

Most of the participants had positive attitude in regard of dental caries affects’ aesthetics and general body health, coherently, studies also showed similar findings [16,13]. Majority of the participants have positive attitude such that tooth decay is avoidable, it is the plausible option for developing nations to prevent than to cure. Moreover, great portion of the subjects said that care of teeth and regular dental checkup is necessary, however, the practice revealed contrary to this. Overall, 73% of the participants had positive attitude. This was comparable score with study done by Pragya.p et.al, in which positive attitude was found in 78.9% of the participants [16].

Although majority of the study participants understand the use of fluoride in toothpaste only 33% use tooth brush with toothpaste. It is a long standing culture in the region to use traditional stick as a method of teeth cleaning, this and availability of the trees can make the students to lean towards it. This was in contradiction with study done by NSHU.B et.al in India, in which toothbrush and paste was common [4]. Nowadays, it is recommended by many dentists to clean teeth at least twice a day [19], but our finding was suboptimal in this regard. Only 5% used pea size toothpaste in our study. There was no one who attend regular dental checkup among the participants and only 12.3% used dentist advised tooth brushing. This is alarming that can question the awareness of students in this regard. The overall practice score of participants tells only 8.7% of the participants scored sufficient practice. The ministry education of Eritrea provides toothbrush and toothpaste to every student, although it is not regular, therefore, this effort should be increased so that the needed practice to be attained.

Although, this study was done with all the existed effort it was not without limit. Variables like; parent background, hygiene status at the time of examination and others would give this study greater scope. Students from outside the town would also be beneficial for comparison purpose. Despite this, it has enough amount of participants with fair distribution relative to the population of the town. The students were assessed for their KAP towards oral health and at the end of data collection the necessary advice regarding oral health was given to all participants, this gives this study dual purpose.

## Conclusion

Overall prevalence of dental caries was 65.3% with mean DMFT index and mean significant index (SiC) of 2.03(SD2.3) and 4.57(SD1.9), respectively. Mean DMFT index was higher among the older age group (19-23 age group) and those who use traditional stick method of tooth cleaning. Our study also founds higher consumption of porridge was associated with higher prevalence of dental caries as well as higher mean DMFT index. Multivariate analysis showed that age group of 19-23 had 80% higher number of DMFT than those 12-14 age category (AIRR 1.8 CI 1.1-3). And those who use traditional method of cleaning tooth had 40% higher number of dmft than those who use tooth brush with paste (AIRR 1.40 CI 1.06-1.8). This study shows most of the participants had sufficient knowledge and positive attitude, 61.3%, 73.8% respectively, however, only 8.7% had adequate oral hygiene practice. Males and females had no difference regarding the overall knowledge, attitude or Practice in our study. Students with adequate practice were higher in junior school than high-school (12.8% vs 5.5%). Knowledge, attitude and practice was positively correlated towards each other in our study, though, not strong.

## Data Availability

All data produced in the present study are available upon reasonable request to the authors

## Ethical Approval and Official Permission

Before starting the survey, ethical approval was obtained from the Ministry of health of Eritrea (Eritrean ministry of health research ethics and protocol review committee) and the directorate of education in Nakfa sub zone and official permission was obtained from the authorities (Principal/Director) of the schools included in the study. Informed consent was obtained from participants’ parents prior to data collection. All the study procedures followed the recommendations of the Helsinki Convention.

## Acknowledgements

The authors would like to thank the teachers and administrators of Nakfa Junior school and Smud junior school as well as Tsabra junior and secondary school for their support and collaboration during the study.

## Author contributions

Conceptualization: MTK and MK. Data curation: MTK, MK, GGG, HT and MEG. Formal analysis: MTK, MK, GGG and OOA. Investigation: MTK, MK, GGG, HT, OOA and MEG. Methodology: MTK and MK. Project administration: MTK and MK. Writing—original draft: MTK, MK, GGG, HT, OOA and MEG. Writing—review MTK, MK, GGG, HT, OOA and MEG. All authors read and approved the final manuscript.

## Funding

The study didn’t receive any funding

## Availability of data and materials

The dataset supporting the conclusions of this article is available from the corresponding author on reasonable request.

## Competing interests

The authors have no conflict of interest to declare in this study

## Notes

### Competing Interest Statement

The authors have declared no competing interest.

### Funding Statement

This study did not receive any funding

### Author Declarations

Ethical Approval and Official Permission Before starting the survey, ethical approval was obtained from the Ministry of health of Eritrea (Eritrean ministry of health research ethics and protocol review committee) and the directorate of education in Nakfa sub zone and official permission was obtained from the authorities (Principal/Director) of the schools included in the study. All the study procedures followed the recommendations of the Helsinki Convention.

